# Assessment of spread of SARS-CoV-2 by RT-PCR and concomitant serology in children in a region heavily affected by COVID-19 pandemic

**DOI:** 10.1101/2020.06.12.20129221

**Authors:** Robert Cohen, Camille Jung, Naim Ouldali, Aurélie Sellam, Christophe Batard, Fabienne Cahn-Sellem, Annie Elbez, Alain Wollner, Olivier Romain, François Corrard, Said Aberrane, Nathalie Soismier, Rita Creidy, Mounira Smati-Lafarge, Odile Launay, Stéphane Béchet, Emmanuelle Varon, Corinne Levy

**Author notes:** **Corresponding author:** Dr Corinne Levy, ACTIV, 31, rue Le Corbusier, 94000 Créteil, France. Email address. Alternate corresponding author: Pr Robert Cohen, ACTIV, 31, rue Le Corbusier, 94000 Créteil, France. Email address.

## Abstract

**Background:** Several studies indicated that children seem to be less frequently infected with SARS-CoV-2 and potentially less contagious. To examine the spread of SARS-CoV-2 we combined both RT-PCR testing and serology in children in the most affected region in France, during the COVID-19 epidemic.

**Methods:** From April 14, 2020 to May 12, 2020, we conducted a cross-sectional prospective, multicenter study. Healthy controls and pauci-symptomatic children from birth to age 15 years were enrolled by 27 ambulatory pediatricians. A nasopharyngeal swab was taken for detection of SARS-CoV-2 by RT-PCR and a microsample of blood for micro-method serology.

**Results:** Among the 605 children, 322 (53.2%) were asymptomatic and 283 (46.8%) symptomatic. RT-PCR testing and serology were positive for 11 (1.8%) and 65 (10.7%) of all children, respectively. Only 3 children were RT-PCR–positive without any antibody response have been detected. The frequency of positivity on RT-PCR for SARS-CoV-2 was significantly higher in children with positive serology than those with a negative one (12.3% vs 0.6%, p<0.001). Contact with a person with proven COVID-19 increased the odds of positivity on RT-PCR (OR 7.8, 95% confidence interval [1.5; 40.7]) and serology (15.1 [6.6;34.6]).

**Conclusion:** In area heavily affected by COVID-19, after the peak of the first epidemic wave and during the lockdown, the rate of children with positive SARS-CoV-2 RT-PCR was very low (1.8%), but the rate of positive on serology was higher (10.7%). Most of PCR positive children had at the same time positive serology.

**What is already known on this topic?:**

- As compared with adults, children seem to be less frequently infected with SARS-CoV-2 and potentially less contagious according to several studies.
- Most of the studies were based on RT-PCR SARS-CoV-2 testing, without antibody assays.

**What this study adds?:**

- This study combining RT-PCR and serologic testing, assessed the spread of SARS-CoV-2 infection in children in area heavily affected by COVID-19 pandemic.
- Among a large cohort of children (>600), 11 (1.8%) were positive on RT-PCR for SARS-CoV-2 and 65 (10.7%) were positive on serology.
- The only factor affecting positivity of RT-PCR for SARS-CoV-2 or serology was the household contact COVID-19.

## Introduction

Since the beginning of the COVID-19 pandemic, reports from several countries indicated that the disease was less frequent and less severe in children than adults.^1-3^ Worldwide, the number of confirmed pediatric cases seems relatively low, and they account for less than 1% of hospitalized cases and deaths.^1, 4^ Although most COVID-19 cases in children are not severe, serious COVID-19 illness resulting in hospitalization can occur in this age group, and recently, hyperinflammatory shock, showing features similar to atypical Kawasaki disease were reported in several countries.^5-10^

However, concerns have been raised that children could play an important role in the spread of the disease because community testing has demonstrated a significant number of children with no or subclinical symptoms.^11^ Indeed, if as for influenza, children could be the primary drivers of household SARS-CoV-2 transmission, then a silent spread from children who did not alert anyone to their infection could be a serious driver in the dynamics of the epidemic.^12^ On the basis of this prevailing hypothesis, school closures were implemented almost ubiquitously around the world to try to halt the potential spread of COVID-19 despite early modelling suggesting that this would have less impact than most other non-pharmacological interventions.^13, 14^

However, several studies had already shown that when SARS-CoV-2 infection was suspected (compatible clinical signs, contact with a person with COVID-19), the rate of positivity on RT-PCR for SARS-CoV-2 was lower in children than adults.^14^.^15^ In contrast, in RT-PCR SARS-CoV-2–positive children, the viral load was comparable between children and adults.^16^ Furthermore, one study suggested that children shed infectious SARS-CoV-2.^17^ However, results from a systematic review of household clusters of COVID-19 revealed that only 3/31 clusters were due to a child index case, and a population-based school contact-tracing study found minimal transmission by child or teacher index cases.^18, 19^ Finally, other studies suggested that children were potentially less contagious than adults.^16, 20-22^

Some countries such as South Korea and Iceland have implemented widespread community testing. Both countries found children significantly underrepresented in cases. In Iceland, this was true in targeted testing of high-risk groups as compared with adults (6.7% < 10 years vs 13.7% ≥ 10 years positive), and in (invited) population screening, no child under 10 years old was positive for SARS-CoV-2 as compared with 0.8% in the general population. ^23^

Of note, all these studies were based on RT-PCR testing, but serology diagnosis is also an important tool to understand the spread and burden of COVID-19.^24^ A serology survey tested adolescents in a high school in the north of France, the site of a cluster at the end of February. Of the 242 students tested, 2.7% of children ≤ 14 years old and 40% aged 15-17 years were positive on SARS-CoV-2 serology (IgG), which suggests a difference in susceptibility to SARS-CoV-2 among younger children.^25^

To best approach the spread and dynamics of transmission of SARS-CoV-2 in children at a population level, we combined both RT-PCR testing for SARS-CoV-2 and serology in asymptomatic or pauci-symptomatic children (with mild clinical symptoms) in the Paris area, the most affected region in France, during the COVID-19 epidemic.

## Patients and Methods

### Study population

This was a cross-sectional prospective, multicenter study conducted by the Association Clinique et Thérapeutique Infantile du Val de Marne (ACTIV) network, a research unit expert in epidemiological surveillance and clinical studies in ambulatory pediatric infectious diseases, and the University Intercommunal Créteil Hospital.^26^ Primary care pediatricians (n = 27) took part in the study from April 14, 2020 to May 12, 2020. The strategy of closing schools and the lockdown decided by the French government for the whole country started on March 17 and finished on May 11, 2020.

This study aimed to enroll children from birth to 15 years of age consulting an ambulatory pediatrician and distributed in two groups: asymptomatic and pauci-symptomatic. Asymptomatic children were defined as children without any symptoms or signs suggesting infectious disease during the last 7 days. In this group, we defined two subgroups of children: those who had history of symptoms (fever or respiratory or digestive) between 7 days and 2 months before enrollment, and those without any history if symptoms. Pauci-symptomatic children were defined as those with fever isolated or associated with respiratory signs such as cough, dysphagia, rhinorrhea, diarrhea, vomiting, cutaneous signs, taste loss and/or anosmia during the last 7 days. Children were excluded if the clinical condition at enrollment required transfer to pediatric emergency unit or hospitalization.

After informing the parents of the participating children and obtaining their signed consent, an electronic case report form (eCRF) was completed by the pediatrician to collect socio-demographic data, history, contact with a person with confirmed COVID-19 by RT-PCR SARS-CoV-2, clinical symptoms and signs, and additional positive biological tests. We have also collected suspected COVID-19 contacts, because of the limited availability of testing. Indeed, during the lockdown, the diagnostic RT-PCR SARS-CoV-2 test was mainly available for patients with severe disease and/or healthcare workers, and all symptomatic individuals could not be tested. For all enrolled children, during the same visit, a nasopharyngeal (NP) swab was taken for RT-PCR detection of SARS-CoV-2 and a microsample of blood for micro-method serology.

### Calculation of the number of patients

To have an appropriate proportion of confirmed RT-PCR SARS-CoV-2–positive patients among asymptomatic children and pauci-symptomatic patients, with a 95% confidence interval (CI) of +/-3%, assuming a positivity proportion < 10%, we needed to enroll 300 children per group (asymptomatic and pauci-symptomatic), for 600 patients in total.

### Serological assays

Pediatricians collected fingerstick whole-blood specimens and used the Biosynex COVID-19 BSS test, a rapid chromatographic immunoassay, for qualitive detection of IgG and IgM antibodies to SARS-CoV-2 in blood. This test targeted the spike protein fragment receptor binding domain and was among those approved by the French national health authority.^27^ According to the specifications of the manufacturer, the diagnostic accuracy of the test was sensitivity 91.8% [95% CI 83.8-96.6] and specificity 99.2% [95%CI 97.7-99.8] (https://www.biosynex.com/laboratoires-hopitaux-tests-covid-19/). However, the accuracy of the test was not stratified by age and for patients infected with other Coronavirus (OC43, 229E, NL63, HKU1) no cross-reaction was observed.

Furthermore, assessment by independent investigators confirmed the good diagnostic accuracy of this test among hospital staff with mild disease in eastern France ^28^.

### SARS-CoV-2 RT-PCR methods

The NP specimens were obtained by using the collection system eSwab^TM^ (Minitip size nylon flocked swab placed in 1 mL of modified liquid Amies transport medium, COPAN, Brescia, Italy). They were transported to the centralized microbiology laboratory (CHIC). Before extraction, each sample was inactivated by the addition of 750 µl / ml of STARmag lysis buffer solution (Seegene, South Korea). The RT-PCR for SARS-CoV-2 was performed on the automated Seegene STARlet system®, according to the manufacturer’s instructions using the CE marked AllplexTM 2019-nCoV RT-PCR assay (Seegene, South Korea®) which targets N- (viral nucleocapsid protein) and RdRP-gene (RNA-dependent RNApolymerase), both SARS-CoV-2 specific genes, and the sarbecovirus specific E-gene.

The automated Hamilton STARlet system was used for automated viral RNA extraction using the STARMag 96 Universal Cartridge kit (Seegene, South Korea) and PCR set up. Subsequently, 8 μL of extracted nucleic acids was added to 17 μL of the PCR Master Mix, and amplification and detection were performed on the CFX96^TM^ detection system (Bio-Rad, France) as per manufacturer’s instruction. Ct from FAM (E gene), Cal Red 610 (RDRP gene), Quasar 670 (N gene) and HEX (internal control) were acquired. Before extraction, internal control (10 µl) was added to verify extraction and determine PCR inhibition. Positive (plasmids encoding the three AllplexTM 2019-nCoV assay target sequences) and negative (RNase-free water added to the master mix prior to PCR) controls were included in each run. The cycle threshold values (Ct) were used as indicators of the copy number of SARS-CoV-2 RNA specimens with lower Ct values corresponding to higher viral copy numbers. NP samples were considered positive when a Ct less than 40 was obtained for any gene. Amplification of two or three targets indicated that SARS-CoV-2 RNA was detected, while amplification of only one target indicated a presumptive positive result. In addition, we defined as weakly positive any result with a Ct > 38 and < 40. A sample was considered negative if the internal control was amplified but not the viral target genes. A sample was considered invalid when no amplification was obtained for the internal control.

### Statistical analysis

Data were entered by using the eCRF (PHP/MySQL) and analyzed by using Stata/SE v15 (StataCorp, College Station, TX, USA). Quantitative data were compared by Student *t* test and qualitative data by chi-square or Fisher exact test. We used a logistic regression model for analysis of factors associated with positivity on RT-PCR for SARS-CoV-2 and serology. Variables (age, clinical signs, contact, siblings and daycare attendance modalities) with p < 0.20 on univariate analysis were included in the model, estimating odds ratios (ORs) and 95% CIs. Only significant variables (p<0.05) were kept in the final model. All tests were 2-sided and were considered significant at p<0.05.

### Ethics

The study protocol was approved by an ethics committee (CPP IDF IX no. 08-022). Parents of all infants provided written informed consent. The study was registered at ClinicalTrials.gov NCT04318431.

## Results

From April 14, 2020 to May 12, 2020, 27 ambulatory pediatricians in the Paris area enrolled 605 children: 322 (53.2%) were asymptomatic and 283 (46.8%) pauci-symptomatic. Table 1 presents the characteristics of the enrolled children by group. In the pauci-symptomatic group, the main signs and symptoms were fever (187, 66.3%), cough (143, 50.7%), pharyngitis (143, 50.7%), rhinitis (137, 48.4%), diarrhea (81, 28.7%), cutaneous criteria (64, 23.0%), vomiting (52, 18.8%), taste loss (8, 3.0%) and anosmia (5, 3.3%).

**Table 1.**
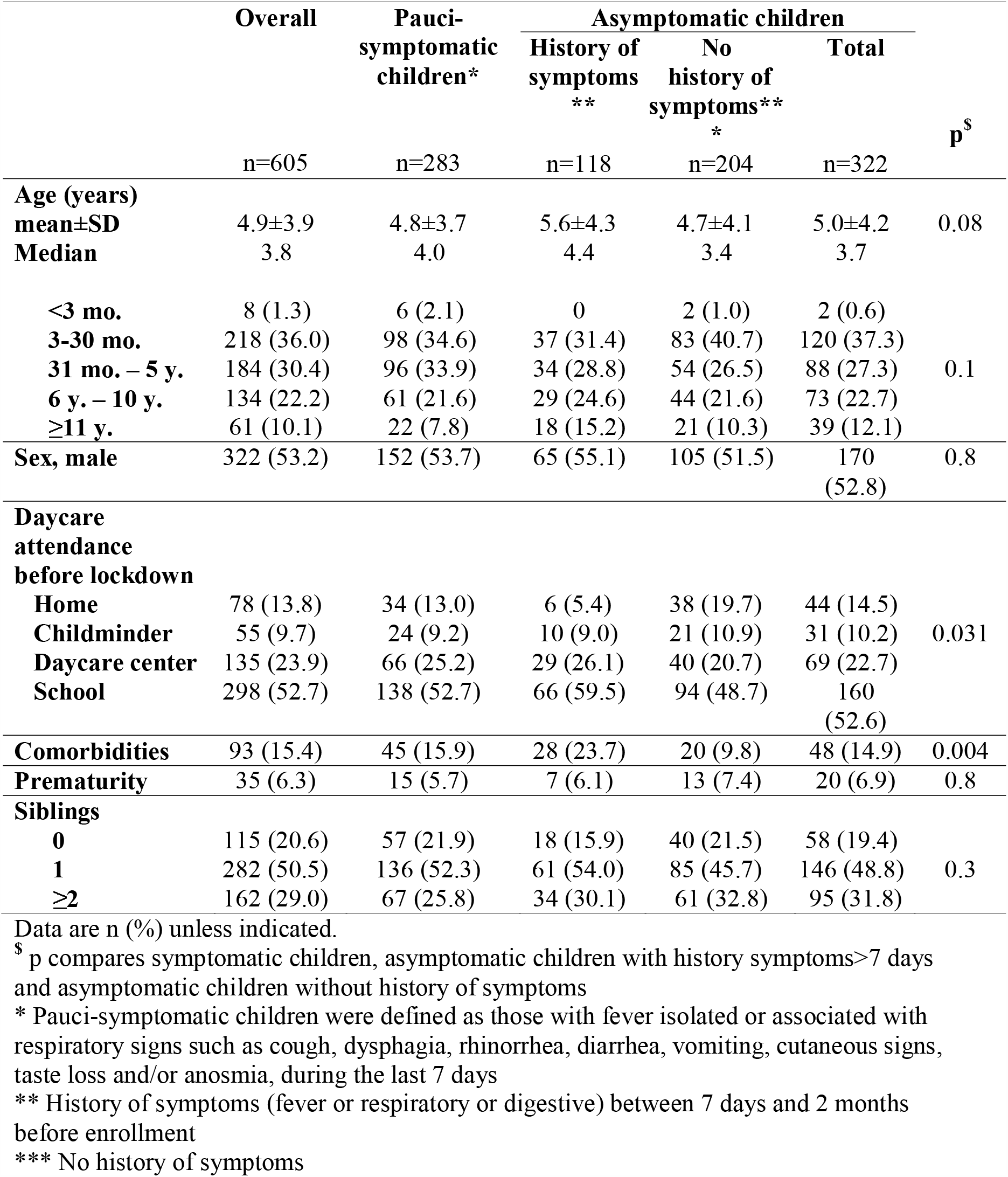
Characteristics of children enrolled in the study and by pauci-symptomatic and asymptomatic group.

| | Overall<br>n=605 | Pauci-symptomatic<br>children*<br>n=283 | Asymptomatic children | | Total<br>n=322 | p<sup>\$</sup> |
| --- | --- | --- | --- | --- | --- | --- |
|  |  |  | History of<br>symptoms<br>**<br>n=118 | No<br>history of<br>symptoms***<br>n=204 |  |  |
| **Age (years)** |  |  |  |  |  |  |
| **mean±SD** | 4.9±3.9 | 4.8±3.7 | 5.6±4.3 | 4.7±4.1 | 5.0±4.2 | 0.08 |
| **Median** | 3.8 | 4.0 | 4.4 | 3.4 | 3.7 |  |
| **<3 mo.** | 8 (1.3) | 6 (2.1) | 0 | 2 (1.0) | 2 (0.6) |  |
| **3-30 mo.** | 218 (36.0) | 98 (34.6) | 37 (31.4) | 83 (40.7) | 120 (37.3) |  |
| **31 mo. – 5 y.** | 184 (30.4) | 96 (33.9) | 34 (28.8) | 54 (26.5) | 88 (27.3) | 0.1 |
| **6 y. – 10 y.** | 134 (22.2) | 61 (21.6) | 29 (24.6) | 44 (21.6) | 73 (22.7) |  |
| **≥11 y.** | 61 (10.1) | 22 (7.8) | 18 (15.2) | 21 (10.3) | 39 (12.1) |  |
| **Sex, male** | 322 (53.2) | 152 (53.7) | 65 (55.1) | 105 (51.5) | 170<br>(52.8) | 0.8 |
| **Daycare**<br>**attendance**<br>**before lockdown** |  |  |  |  |  |  |
| **Home** | 78 (13.8) | 34 (13.0) | 6 (5.4) | 38 (19.7) | 44 (14.5) |  |
| **Childminder** | 55 (9.7) | 24 (9.2) | 10 (9.0) | 21 (10.9) | 31 (10.2) | 0.031 |
| **Daycare center** | 135 (23.9) | 66 (25.2) | 29 (26.1) | 40 (20.7) | 69 (22.7) |  |
| **School** | 298 (52.7) | 138 (52.7) | 66 (59.5) | 94 (48.7) | 160<br>(52.6) |  |
| **Comorbidities** | 93 (15.4) | 45 (15.9) | 28 (23.7) | 20 (9.8) | 48 (14.9) | 0.004 |
| **Prematurity** | 35 (6.3) | 15 (5.7) | 7 (6.1) | 13 (7.4) | 20 (6.9) | 0.8 |
| **Siblings** |  |  |  |  |  |  |
| **0** | 115 (20.6) | 57 (21.9) | 18 (15.9) | 40 (21.5) | 58 (19.4) |  |
| **1** | 282 (50.5) | 136 (52.3) | 61 (54.0) | 85 (45.7) | 146 (48.8) | 0.3 |
| **≥2** | 162 (29.0) | 67 (25.8) | 34 (30.1) | 61 (32.8) | 95 (31.8) |  |

Data are n (%) unless indicated.
<sup>\$</sup> p compares symptomatic children, asymptomatic children with history symptoms>7 days and asymptomatic children without history of symptoms
\* Pauci-symptomatic children were defined as those with fever isolated or associated with respiratory signs such as cough, dysphagia, rhinorrhea, diarrhea, vomiting, cutaneous signs, taste loss and/or anosmia, during the last 7 days
\*\* History of symptoms (fever or respiratory or digestive) between 7 days and 2 months before enrollment
\*\*\* No history of symptoms

Figure 1 presents the dynamics of the first epidemic wave in Paris area^29^, the dates of the lockdown and the number of children enrolled by weeks.

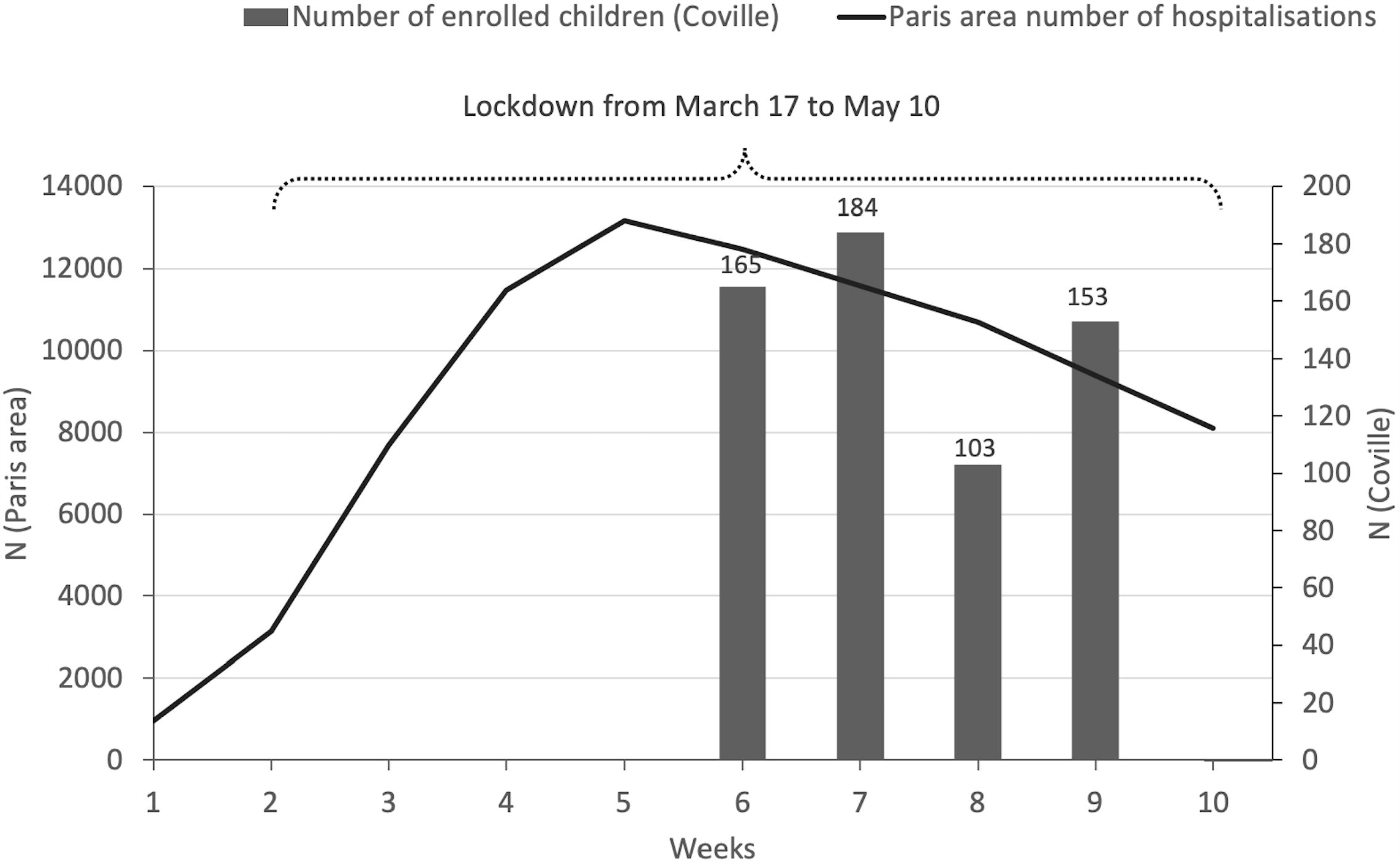

RT-PCR SARS-CoV-2 tests were positive for 11 (1.8%) children, with no significant difference between the two groups (Table 2). The supplemental Table shows the details of the 11 positive RT-PCR for SARS-CoV-2. Only 3 children had positive RT-PCR SARS-CoV-2 result with Ct less than 31.

**Table 2.** Results of RT-PCR SARS-Cov-2 testing and serology in children by pauci-symptomatic and asymptomatic group.

|  | Overall | Pauci-symptomatic children* | Asymptomatic children |  |  |
| --- | --- | --- | --- | --- | --- |
|  |  |  | History of symptoms** | No history of symptoms** | Total |
|  | n=605 | n=283 | n=118 | n=204 | n=322 |
| **RT-PCR** |  |  |  |  |  |
| **Overall** | 11 (1.8)<br>[0.9; 3.2] | 7 (2.5)<br>[1.0; 5.0] | 1 (0.8)<br>[0.0; 4.6] | 3 (1.5)<br>[0.3; 4.2] | 4 (1.2)<br>[0.3; 3.1] |
| **Definite positive** | 5 | 3 | 0 | 2 | 2 |
| **Weakly positive** | 1 | 1 | 0 | 0 | 0 |
| **Probable** | 5 | 3 | 1 | 1 | 2 |
| **Serology** |  |  |  |  |  |
| **IgM+ and/or IgG+** | 65 (10.7)<br>[8.4; 13.5] | 24 (8.5)<br>[5.5; 12.4] | 28 (23.7)<sup>\$</sup><br>[16.4; 32.4] | 13 (6.4)<sup>\$</sup><br>[3.4; 10.7] | 41 (12.7)<br>[9.3; 16.9] |
| **IgM+IgG-** | 7 (1.2)<br>[0.5; 2.4] | 4 (1.4)<br>[0.4; 3.6] | 2 (1.7)<br>[0.2; 6.0] | 1 (0.5)<br>[0.0; 2.7] | 3 (0.9)<br>[0.2; 2.7] |
| **IgM+IgG+** | 32 (5.3)<br>[3.6; 7.4] | 12 (4.2)<br>[2.2; 7.3] | 17 (14.4)<sup>\$</sup><br>[8.6; 22.1] | 3 (1.5)<sup>\$</sup><br>[0.3; 4.2] | 20 (6.2)<br>[3.8; 9.4] |
| **IgM-IgG+** | 26 (4.2)<br>[2.8; 6.2] | 8 (2.8)<br>[1.2; 5.5] | 9 (7.6)<br>[3.5; 14.0] | 9 (4.4)<br>[2.0; 8.2] | 18 (5.6)<br>[3.3; 8.7] |

Data are n (%) [95% confidence interval].
<sup>\$</sup> p<0.001
\* Pauci-symptomatic children were defined as those with fever isolated or associated with respiratory signs such as cough, dysphagia, rhinorrhea, diarrhea, vomiting, cutaneous signs, taste loss and/or anosmia, during the last 7 days
\*\* History of symptoms (fever or respiratory or digestive) between 7 days and 2 months before enrollment
\*\*\* No history of symptoms

On multivariable analysis, contact with a person with proven COVID-19 was the only significant risk factor for RT-PCR–positive SARS-CoV-2 infection (OR 7.8, 95% CI [1.5; 40.7]).

Table 2 shows also the serology results by group. Serology was positive for 65 (10.7%) children, whatever the group, and among these, 87.3% had a confirmed or suspected contact. Children with history of symptoms during the preceding weeks, more frequently were positive on serology.

Table 3 presents the RT-PCR SARS-CoV-2 results by serology status. The frequency of positivity was significantly higher in children with positive serology than those with a negative one (12.3% vs 0.6%, p<0.001). Only 3 children were RT-PCR SARS-CoV-2– positive without any antibody response detected.

**Table 3.** RT-PCR SARS-CoV-2–positive results by serology status.

| **RT-PCR results** | **IgM-**<br>**IgG-**<br>**n=540** | **IgM+**<br>**and/or IgG+**<br>**n=65** | **IgM+**<br>**IgG-**<br>**n = 7** | **IgM+**<br>**IgG+**<br>**n = 32** | **IgM -**<br>**IgG+**<br>**n = 26** |
| --- | --- | --- | --- | --- | --- |
| Overall positive* | 3 (0.6) | 8 (12.3) | 0 | 6 (18.7) | 2 (7.8) |
| Definite positive** | 2 (0.4) | 3 (0.6) | 0 | 2 (6.2) | 1 (3.9) |
| Weakly positive *** | 0 | 1 (1.5) | 0 | 1 (3.1) | 0 |
| Presumptive positive | 1 (0.2) | 4 (6.2) | 0 | 3 (9.4) | 1 (3.9) |

\*\*\*\*
Data are n (%).
- p<0.001: comparison of overall RT-PCR with IgM-/IgG- vs IgM+ and/or IgG+
- p<0.001: comparison of definite or weakly or probable RT-PCR with IgM-/IgG- vs IgM+ and/or IgG+
\* Overall positive: NP samples were considered positive when a cycle threshold value (Ct) less than 40 was obtained for any gene
\*\* Definite positive: cycle threshold value (Ct) less than 38 obtained for 2 or 3 gene.
\*\*\* Weakly positive: any result with a Ct > 38 and < 40.
\*\*\*\* Presumptive: cycle threshold value (Ct) less than 38 obtained for only one target.

Table 4 shows RT-PCR SARS-CoV-2 and serology positivity by contact with a person with suspected or confirmed COVID-19. Only 2 of 275 (0.7%) children without any contact with a person with COVID-19 were positive on RT-PCR for SARS-CoV-2.

**Table 4.** RT-PCR and serology results by contact with a person with confirmed and/or suspected COVID-19.

| **Contact** | **Overall**<br>**n=543\*** | **Positive**<br>**serology**<br>**n=63** | **Negative**<br>**serology**<br>**n=480** | **Positive**<br>**RT-PCR**<br>**SARS-CoV-2**<br>**n=11** | **Negative**<br>**RT-PCR SARS-**<br>**CoV-2**<br>**n=532** |
| --- | --- | --- | --- | --- | --- |
| Confirmed<br>COVID-19** | 93 (17.1)<br>[14.1; 20.6] | 29 (31.2)<br>[22.0; 41.6] | 64 (68.8)<br>[58.4; 78.0] | 5 (5.4)<br>[1.8; 12.1] | 88 (94.6)<br>[87.9; 98.2] |
| Suspected<br>COVID-19*** | 175 (32.2)<br>[28.3; 36.3] | 26 (14.9)<br>[9.9; 21.0] | 149 (85.1)<br>[79.0; 90.0] | 4 (2.3)<br>[0.6; 5.7] | 171 (97.7)<br>[94.3; 99.4] |
| Confirmed/<br>suspected<br>COVID-19 | 268 (49.4)<br>[45.1; 53.6] | 55 (20.5)<br>[15.9; 25.9] | 213 (79.5)<br>[74.1; 84.1] | 9 (3.4)<br>[1.5; 6.3] | 259 (96.6)<br>[93.7; 98.5] |
| No contact | 275 (50.6)<br>[46.4; 54.9] | 8 (2.9)<br>[1.3; 5.7] | 267 (97.1)<br>[94.3; 98.7] | 2 (0.7)<br>[0.1; 2.6] | 273 (99.3)<br>[97.4; 99.9] |

Data are n (%) [95% confidence interval].
\* 543 available data on 605 enrolled patients
\*\* Confirmed by RT-PCR SARS-CoV-2
\*\*\* Suspected symptoms suggestive of COVID-19 because of the limited availability of testing

On multivariate analysis, positivity on serology was associated with contact with a person with proven or suspected COVID-19 (OR 15.1 [95% CI 6.6; 34.6] and 5.8 [95% CI 2.6; 13.2]).

## Discussion

This study combines RT-PCR SARS-CoV-2 and serology results to assess the spread of SARS-CoV-2 infection in a large cohort of children in the community. In a region strongly affected by the epidemic (Paris area), but during the lockdown, very few children (1.8%) were positive on RT-PCR for SARS-CoV-2, but the rate of children positive on serology (10.7%) was higher. Despite the relatively large number of children included (>600), we did not find a significant difference in the rate of positive RT-PCR or serology results between asymptomatic and pauci-symptomatic children.

Among asymptomatic children, those with no history of symptoms during the preceding weeks accounted for two third of children with positive serology results (28/41), which supports that asymptomatic infections are frequent in children. By contrast, history of symptoms during the preceding weeks increased significantly the risk of positive serology. However, on multivariate analysis, the only factor influencing the positivity of RT-PCR or serology was the household contact who has previously presented symptoms suggestive of COVID-19. Of note, the number of siblings in the family did not significantly increase the probability of a positive RT-PCR or serology result. Several studies have shown that children were usually infected by an adult in the family.^18, 22, 30, 31^ In our study, the importance of familial contagion in the modalities of SARS-Cov-2 transmission is suggested by a very low RT-PCR (0.7%) and serology positivity rate (3.6%) for children without an infected relative and in a period of lockdown.

Among the children positive on RT-PCR (n=11), only 3 had no antibody response, and 8 were positive for IgG with or without IgM positivity. This finding supports that for these 3 patients, contamination had occurred during the 2 weeks before enrollment.

We highlight that the frequency of positivity on RT-PCR for SARS-CoV-2 was significantly higher in children with positive serology than those with a negative one (12.3% vs 0.6%, p<0.001). This finding highlights the difficulties in interpreting the significance of a positive RT-PCR SARS-CoV-2 result without concomitant antibody testing after the epidemic wave. Indeed, children positive on RT-PCR for SARS-CoV-2 and positive for IgG probably had little or no infectivity.^32^ In a study of 9 patients, attempts to isolate the virus in culture were not successful beyond day 8 of illness onset, which relates to the decreased infectivity beyond the first week.^33^ In the study of Bullard *et al*., SARS-CoV-2 Vero cell infectivity was only observed for RT-PCR Ct < 24 and symptom onset to test < 8 days.^34^ It is likely that infectivity was low for the 8 of 11 RT-PCR SARS-CoV-2 positive children. Indeed, only 3 children had a Ct recorded under 31.

Our study has several limitations. First, the role of assumed household transmission probably has been over-estimated because of the well-followed lockdown in France.^35^ Indeed, more than 86.5% of children with positive SARS-CoV-2 by RT-PCR or serology have had a confirmed or suspected COVID-19 household contact. However, our rate of positive serology for children in the Paris area was similar to the rate observed for hospitalized patients (11.7%) and at school children (8.8%).^22, 36^ Second, the ability to successfully collect nasopharyngeal swabs properly could be more difficult in young children and significantly affect the results and be a factor contributing to the low RT-PCR positivity prevalence observed in our population. However, the pediatricians who performed the study were all involved for many years in a pneumococcal nasopharyngeal carriage study (started in 2001 and currently ongoing) and were particularly well trained to collect appropriately nasopharyngeal samples.^37^

School closure or limitation (reduced number of students or days of attendance) has a major impact on children’s development and access to learning.^38^ Therefore, the usefulness of school closure or limitation needs evaluation in controlling the COVID-19 epidemic. ^39^ We plan to renew this study after the full re-opening of schools and day care centre in the Paris area to better assess the transmission of SARS-Cov-2 in children.

## Data Availability

Data are available upon reasonable request

## Acknowledgments

We are grateful to the investigators of the COVILLE study Network:

Akou’ou M.H, Auvrignon A, Belaroussi N, Benani M, Cambier Nappo E, Chartier Albrech C, Coicadan L, Condor R, D’Acremont G, D’Ovidio N, De Brito B, Deberdt P, Delatour A, Gorde-Grosjean S, Louvel M, Michot-Cottias A-S, Ravilly S, Seror E, Turberg-Romain C, We are grateful to Adjemian S, Auffroy O, Begard M, Harant J, Mouaouya M, Receveau F, and Sigere ML, for technical assistance ; Cuquemelle A for secretarial assistance.

We are grateful to the ACTIV team : Ramay I, Prieur C; Prieur A, Borg M, Meyet L, Levy J, and Zemmour E (Association Clinique et thérapeutique Infantile du Val de Marne);

We are grateful to the CRC team: Brussieux M and Hoffart C from the Clinical Reaserch Center of the CHI Créteil.

